# Population-level relative effectiveness of the COVID-19 vaccines and the contribution of naturally acquired immunity

**DOI:** 10.1101/2022.10.04.22280689

**Authors:** Kayoko Shioda, Yangping Chen, Matthew H Collins, Benjamin A Lopman

## Abstract

**Background:** Immune protection against SARS-CoV-2 can be induced by natural infection or vaccination or both. The interaction between vaccine-induced immunity and naturally acquired immunity at the population level has been understudied.

**Methods:** We used regression models to evaluate whether the impact of COVID-19 vaccines differed across states with different levels of naturally acquired immunity from March 2021 to April 2022 in the United States. Analysis was conducted for three evaluation periods separately (Alpha, Delta, and Omicron waves). As a proxy of the proportion of the population with naturally acquired immunity, we used either the reported seroprevalence or the estimated proportion of the population ever infected in each state.

**Results:** COVID-19 mortality decreased as the coverage of ≥1 dose increased among people ≥65 years of age, and this effect did not vary by seroprevalence or the proportion of the total population ever infected. Seroprevalence and the proportion ever infected were not associated with COVID-19 mortality, after controlling for vaccine coverage. These findings were consistent in all evaluation periods.

**Conclusions:** COVID-19 vaccination was associated with a sustained reduction in mortality at the state level during the Alpha, Delta, and Omicron periods. The effect did not vary by naturally acquired immunity.

## INTRODUCTION

Immune protection against SARS-CoV-2 can be induced by natural infection or vaccination or both. The understanding of the relationship between immune responses following natural infection and vaccination is constantly advancing, and becomes further complicated as different variants of concern have caused multiple waves of infection over the course of the pandemic. Heterogeneity in both the prevalence of infection and vaccine coverage has been observed over time in the United States, providing an opportunity to evaluate the interaction between naturally acquired immunity and vaccine-induced immunity at the population level.

We asked whether vaccine effectiveness at the population level would differ across states with different proportions of the population with naturally acquired immunity. According to the Centers for Disease Control and Prevention (CDC), the seroprevalence was highest in Wisconsin (24.0%) and lowest in Vermont (1.4%) at the time of vaccine rollout (December 2020) [1]. Thus, some states vaccinated a partially immune population, while other states vaccinated an almost completely immunologically naive population. We hypothesized that the population-level impact of vaccination would be greatest among states with higher rates of previous SARS-CoV-2 infections for at least two reasons: 1) Vaccinating those with prior infection leads to hybrid immunity, which may be more robust and durable; and 2) Populations with higher proportions of naturally immune may more quickly reach a level of herd immunity that diminishes transmission, thus limiting infection and disease by reducing exposure.

Since the vaccine became available to the general public in the U.S., the coverage considerably differed across states. As of May 24, 2022, the percentage of the population fully vaccinated is highest in Rhode Island (83.1%) and lowest in Wyoming (51.1%) [1]. Alongside vaccine administration, the country continued to experience multiple waves of different variants, infecting many people before and/or after vaccination. People who received vaccination after natural infection were found to have higher levels of neutralizing antibodies (NAb) when compared to vaccinated individuals without previous infection [2,3]. Those who got infected after vaccination (i.e., breakthrough infection) were also found to have higher NAb level compared to those who only had vaccination [4]. This becomes more complicated as different variants of concern have swept through the world, exposing the population to heterologous spike proteins. Immune imprinting from initial antigen exposure plays an important role and alters immune response after vaccination or natural infection [5–7]. For example, vaccine responses among people with previous infection were found to be less effective if the previous infection was caused by a variant with significantly different spike sequences [5].

While individual-level findings on the relationship between naturally acquired immunity and vaccine-induced immuntiy have been evolving, we do not yet know whether the population-level COVID-19 vaccine impact differs by the proportion of the population that had immunity from natural infection, and if so, how. Therefore, we evaluated how the relationship between COVID-19-related deaths and vaccine coverage differed across states with different levels of naturally acquired immunity. Three periods were analyzed separately: the Alpha wave, the Delta wave, and the Omicron wave. We used two approaches to define the proportion of a state’s population with naturally-acquired immunity. First, we used state-level seroprevalence reported by the CDC, assuming that antibodies and immune protection wane at a similar speed [1]. Second, we estimated the proportion of the population ever infected based on serosurvey data [8] and also used estimates from other studies [9,10].

## METHODS

### Overall study design and evaluation periods

We evaluated whether the relationship between the number of COVID-19 deaths and COVID-19 vaccine coverage varied by the reported state-level seroprevalence (or the estimated proportion ever infected) during each of the following three evaluation periods: Alpha wave (from March 1 to June 30, 2021), Delta wave (from July 1 to December 15, 2021), and Omicron wave (from December 16, 2021 to April 11, 2022) in the U.S. We analyzed these evaluation periods separately because each SARS-CoV-2 variant may exhibit unique transmissibility and pathogenicity.

### State-level COVID-19 data

For state-level COVID-19 mortality data, we used the publicly available daily time series data downloaded from the CDC website on April 21, 2022 [11]. The date of death was from January 22, 2020 to April 19, 2022. The number of COVID-19 deaths per one million population was calculated using the U.S. census population data for each state [12]. We calculated the 7-day average of the daily death counts and created weekly times series by summing the average daily counts in each week. Negative weekly death counts, which were observed in nine weeks in six states, were set to zero.

For state-level seroprevalence, we used the publicly available CDC seroprevalence data downloaded on March 17, 2022 [13]. Details of this seroprevalence data can be found elsewhere [14,15], but briefly, the survey has collected convenience samples of deidentified residual patient sera in 10 U.S. sites from March to July 2020 and in all 50 states from August 2020 to present. The serosurvey round used for each of the evaluation periods was Round 15 (samples collected between February and early March) for the Alpha wave, Round 24 (between late June and early July) for the Delta wave, and Round 28 (between end of November and mid/late December) for the Omicron wave.

For state-level vaccine coverage, the publicly available daily time series data on the proportions of the population that received ≥1 dose, full doses, and a booster dose were downloaded from the CDC website on April 22, 2022 [11]. The data were stratified by age group. The date of vaccine administration was from December 13, 2020 to April 20, 2022.

### Regression analysis

We ran a regression analysis to evaluate the variation in the population-level impact of COVID-19 vaccines against mortality by the proportion of the population with naturally acquired immunity in each state. All analyses were conducted with R (R Center for Statistical Computing; Vienna, Austria) v4.

For the Alpha wave, we fit a mixed-effect linear regression model with random intercepts and slopes for each state. An outcome variable was weekly COVID-19 death counts per 1 million population in each state. Independent variables were the reported CDC seroprevalence among people ≥65 years of age at the beginning of the Alpha wave in each state, weely proportion of people ≥65 years of age who had ≥1 dose of COVID-19 vaccines in each state, and an interaction term of these two variables. The nlme R software package was used to fit this mixed-effect model [16]. In the sub-analysis for the Alpha wave, we incorporated a lag between infection and death (3 weeks) and a lag between vaccination and immunity acquisition (2 weeks) in the regression model.

For the Delta wave and Omicron wave, we fit linear regression models using the total number of deaths per 1 million population during each wave in each state as an outcome. Independent variables were the state-level seroprevalence among people ≥65 years of age at the beginning of each wave, the average proportion of people ≥65 years of age who had ≥1 dose of COVID-19 vaccines during each wave in each state, and an interaction of these two variables.

### Sub-analysis

We used different variables to define the proportion of the population with naturally acquired immunity in the sub-analysis. For the Alpha wave, we used the proportion of the total population ever infected in each state before vaccine introduction, which was estimated using the previously developed Bayesian model [8] (see the next section “Estimating the proportion of the population ever infected” for more information). We also used estimates of the proportion of the total population ever infected as of December 2020 in each state [9]. For the Omicron wave, we used the proportion of the total population ever infected as of November 14, 2022 [10].

In addition, for vaccine coverage, we used the average proportions of people ≥65 years of age who were fully vaccinated and who received a booster dose in each state in the sub-analysis, instead of the proportion with ≥1 dose that was used in the main analysis.

### Estimating the proportion of the population ever infected

We estimated the proportion of the population ever infected in 50 states as of December 2020 (around the time of COVID-19 vaccine introduction in the U.S.) based on state-level cross-sectional seroprevalence [8]. We adjusted seroprevalence for the timeline of antibody waning, given the considerable evidence that antibodies against SARS-CoV-2 wane below a detectable level over time [17–20]. We first determined the timing of symptom onset for fatal COVID-19 cases based on empirical data on the number of days between symptom onset and death [8].

We then used a Markov chain Monte Carlo model to estimate the mean of the Weibull distribution for time of seropositivity (i.e., time from acquisition to loss of the detectable level of antibodies) and infection fatality ratio (IFR) based on daily time series data for COVID-19 mortality and repeated cross-sectional seroprevalence data in each state (see “State-level COVID-19 data” above). The estimated number of infections on each day was calculated by adjusting the number of reported deaths for the estimated IFR and the lag between infection and death. North Dakota was excluded due to the limited seroprevalence data to fit the model.

## RESULTS

### Seroprevalence and vaccine coverage

The median seroprevalence among people aged ≥65 years at the beginning of each evaluation period was 12.9%, 11.3%, and 19.6% in the Alpha wave, the Delta wave, and the Omicron wave in the U.S., respectively (Table 1). A great variation in the reported seroprevalence was observed among this age group in each wave. The reported seroprevalence was lowest in Hawaii (1.0%) and highest in Wisconsin (26.4%) in the Alpha wave, lowest in Vermont (0%) and highest in Utah (24.4%) during the Delta wave, and lowest in Hawaii (5.8%) and highest in Wisconsin (29.1%) during the Omicron wave.

**Table 1.** Descriptive statistics of the state-level SARS-CoV-2 seroprevalence, COVID-19 vaccine coverage, and COVID-19 mortality during the Alpha, Delta, and Omicron wave.

|  | Median (min, max) |  |  |
| --- | --- | --- | --- |
|  | Alpha wave | Delta wave | Omicron wave |
| CDC serorevalence among people ≥65 years of age at the beginning of each evaluation period | 12.9%<br>(1.0%, 26.4%) | 11.3%<br>(0%, 24.4%) | 19.6%<br>(5.8%, 29.1%) |
| Average percentage of people ≥65 years of age that received at least one dose of COVID-19 vaccines | 77.4%<br>(67.6%, 88.9%) | 91.9%<br>(82.1%, 99.6%) | 95.0%<br>(90.7%, 95.0%) |
| Average percentage of people ≥65 years of age that received full doses of COVID-19 vaccines | 62.6%<br>(49.6%, 72.7%) | 83.5%<br>(72.4%, 95.8%) | 89.2%<br>(79.2%, 95.0%) |
| Average percentage of people ≥65 years of age that received a booster dose of COVID-19 vaccines | Not available | 14.4%<br>(3.3%, 19.3%) | 65.2%<br>(29.5%, 79.7%) |
| Total deaths per 1 million (all age) | 192.2<br>(56.2, 443.2) | 568.1<br>(91.7, 1314.4) | 520.1<br>(180.2, 973.6) |

The coverage of ≥1 dose of COVID-19 vaccines among people who are ≥65 years of age increased over time, with more variability closer to the time of vaccine introduction. The coverage of ≥1 dose reached a high level (>90%) in all states by the Omicron wave. The average coverage of the booster dose in this age group had a large variation in the Omicron wave; New Hampshire had the lowest (29.5%) and Minnesota had the highest (79.7%).

### Regression analysis

Our regression analysis found three trends in the relationship of COVID-19 mortality with vaccine coverage and the proportion of the population with naturally acquired immunity, which were consistently found in all three evaluation periods. First, COVID-19 mortality decreased as vaccine coverage among people aged 65 years and older increased, after controlling for the seroprevalence. In the Alpha wave, the average weekly death counts was 9.2 per 1 million. Per a 10% increase in weekly coverage of ≥1 dose among people ≥65 years of age, weekly death counts decreased by 2.9 (95% confidence interval (CI): 2.2-3.7) per 1 million population during this evaluation period (Supplementary Table 1). The same trend was observed after incorporating the lag between infection and death and the lag between vaccine administration and immunity acquisition (Supplementary Table 2). Per a 10% increase in the coverage of ≥1 dose among people ≥65 years of age, total COVID-19 deaths during each wave was associated with a decrease of 420 (95% CI: 273-567. Supplementary Table 3) and 443 (5-882. Supplementary Table 4) per 1 million population in the Delta wave and Omicron wave, respectively.

Second, the seroprevalence among people ≥65 years of age was not associated with changes in COVID-19 mortality, after controlling for vaccine coverage among people ≥65 years of age (Supplementary Table 1-4). Third, the degree of decrease in COVID-19 mortality per a unit increase in vaccine coverage among people ≥65 years of age did not vary by state-level seroprevalence among people aged ≥65 years in all evaluation periods. In all models, 95% CIs for the interaction terms between the vaccine coverage and seroprevalence included the null value (Supplementary Table 1-4).

### Sub-analysis using the estimated proportion of the population ever infected

To define the proportion of the population with naturally acquired immunity differently, we estimated the proportion of the total population ever infected and IFR by December 2020 in each state by adjusting the state-level seroprevalence for the timeline of waning antibodies (Table 2). The average duration of seropositivity (as an indicator of waning) was estimated to be 160 days for New York and 192 days for Pennsylvania at the population level. We could not estimate the average duration of seropositivity in other states, either because a peak in the seroprevalence likely happened before the beginning of national serosurveillance in August 2020 or the decline in seroprevalence was not observed. In these remaining states, we ran the model using the average of the estimated duration of seropositivity in New York and Pennsylvania. The model did not fit the observed seroprevalence data in five states (Connecticut, the District of Columbia, Massachusetts, New Jersey, and, Rhode Island) which were excluded from the following analysis.

**Table 2.** Estimated state-level proportions of the population ever infected and infection fatality ratios as of December 2020 in the U.S.

| State | Estimated average IFR | % ever infected as of Dec 31, 2020 |
| --- | --- | --- |
| AK | 0.27 (0.25-0.29) | 12.1% (11.1-13.2%) |
| AL | 0.55 (0.52-0.58) | 24.1% (22.9-25.3%) |
| AR | 0.87 (0.82-0.92) | 16.6% (15.7-17.6%) |
| AZ | 0.87 (0.83-0.92) | 18.8% (17.8-19.7%) |
| CA | 0.54 (0.51-0.58) | 16.2% (15.3-17.3%) |
| CO | 0.6 (0.54-0.66) | 16% (14.4-17.6%) |
| DE | 0.66 (0.62-0.7) | 16.9% (15.9-17.9%) |
| FL | 0.73 (0.69-0.77) | 16.6% (15.7-17.5%) |
| GA | 0.55 (0.53-0.58) | 22.6% (21.5-23.7%) |
| HI | 0.52 (0.41-0.67) | 4.6% (3.6-5.8%) |
| IA | 0.45 (0.43-0.48) | 31% (29.5-32.5%) |
| ID | 0.56 (0.5-0.64) | 16.7% (14.8-18.9%) |
| IL | 0.48 (0.46-0.5) | 32% (30.8-33.3%) |
| IN | 1.36 (1.13-1.65) | 10.7% (8.8-12.9%) |
| KS | 0.64 (0.6-0.68) | 18.7% (17.5-19.9%) |
| KY | 0.48 (0.45-0.51) | 15.5% (14.6-16.5%) |
| LA | 0.86 (0.82-0.91) | 20.5% (19.5-21.6%) |
| MD | 0.49 (0.47-0.52) | 22.3% (21.1-23.5%) |
| ME | 1.2 (0.98-1.51) | 3% (2.4-3.7%) |
| MI | 0.73 (0.68-0.77) | 20% (18.8-21.5%) |
| MN | 0.47 (0.45-0.5) | 22.3% (21.2-23.5%) |
| MO | 0.52 (0.49-0.55) | 20.1% (19.1-21.1%) |
| MS | 0.78 (0.74-0.81) | 23.7% (22.6-24.9%) |
| MT | 0.67 (0.6-0.74) | 16% (14.4-17.7%) |
| NC | 0.58 (0.54-0.62) | 14.1% (13.2-15%) |
| NE | 0.31 (0.3-0.33) | 30.5% (29.2-31.9%) |
| NH | 1.69 (1.5-1.93) | 4.1% (3.6-4.6%) |
| NM | 0.69 (0.65-0.73) | 20.4% (19.3-21.6%) |
| NV | 0.55 (0.52-0.58) | 23.6% (22.5-24.7%) |
| NY | 0.44 (0.43-0.46) | 37.9% (36.4-39.5%) |
| OH | 0.54(0.52-0.57) | 25% (23.8-26.3%) |
| OK | 0.5 (0.47-0.53) | 16% (14.9-17%) |
| OR | 0.55 (0.5-0.61) | 7.7% (7-8.5%) |
| PA | 0.6 (0.57-0.63) | 25.1% (23.9-26.3%) |
| SC | 0.75 (0.71-0.79) | 17.1% (16.2-18.1%) |
| SD | 0.75 (0.61-0.99) | 25.1% (19.1-31.1%) |
| TN | 0.5 (0.48-0.53) | 25.6% (24.4-26.9%) |
| TX | 0.51 (0.48-0.53) | 23.1% (22-24.2%) |
| UT | 0.21 (0.19-0.23) | 23.9% (21.8-25.9%) |
| VA | 0.74 (0.68-0.8) | 9.7% (9-10.5%) |
| VT | 1.07 (0.78-1.52) | 2.4% (1.7-3.3%) |
| WA | 0.69 (0.63-0.76) | 7.8% (7.1-8.6%) |
| WI | 0.35 (0.33-0.37) | 29.2% (27.8-30.7%) |
| WV | 0.92 (0.84-1.02) | 10.3% (9.3-11.3%) |
| WY | 0.4 (0.35-0.45) | 22.9% (20-26.2%) |

Abbreviations: IFR, infection fatality ratio.
Five states (CT, DC, MA, NJ, RI) were excluded because of the bad fit to the data. One state (ND) was excluded due to the limited seroprevalence data to fit the model to.

When using the estimated proportions of the total population ever infected as of December 2020 instead of the seroprevalence to define the proportion of the population with naturally acquired immunity for the Alpha wave, the overall findings remained the same (Supplementary Table 5). The coverage of ≥1 dose of COVID-19 vaccines was associated with a decline in COVID-19 mortality at the state level, while the effect did not vary by the estimated proportion of the population ever infected in the Alpha wave. We also used other estimates of the proportion of the population ever infected for the Alpha wave [9] and Omicron wave [10] and found the same trends (Supplementary Table 6 and 7).

### Sub-analysis using other measures of vaccine coverage

In the Delta wave, we found the same results after using the proportion of people aged ≥65 years that were fully vaccinated, instead of ≥1 dose; Total COVID-19 deaths decreased as the coverage increased, while the seroprevalence or the proportion ever infected did not affect COVID-19 mortality, after controlling for vaccine coverage. For the Omicron wave, the results remained consistent when using the proportion fully vaccinated instead of the proportion with ≥1 dose. However, when we used the proportion of people ≥65 years of age that received a booster to define the vaccine coverage, there was no association between COVID-19 mortality and booster coverage.

## DISCUSSION

We assessed the relationship between vaccine-induced immunity and naturally acquired immunity against COVID-19 mortality at the population level in the U.S. in 2021-2022. COVID-19 death counts decreased as the coverage of ≥1 dose increased among ≥65 years of age, and this effect did not vary by seroprevalence or the proportion of the population ever infected. Seroprevalence and the proportion ever infected were not associated with COVID-19 mortality, after controlling for vaccine coverage. These findings were consistent in all three evaluation periods. These results indicate that vaccine coverage is associated with protection against deaths that is visible at the population level, compared to naturally acquired immunity. This suggests that we should encourage people to receive vaccination, especially primary doses, to prevent severe outcomes, instead of relying on immunity from natural infection.

The population-level impact of COVID-19 vaccines did not vary by the seroprevalence and the estimated proportion of the population ever infected in all evaluation periods. This might be because the people who got infected and the people who received vaccines were different, especially in the Alpha period, making it hard to see a combined effect. In the Delta and Omicron waves, both the vaccine coverage and seroprevalence reached high levels, so there was likely a good amount of overlap between these two groups. However, we still did not see a varied effect of vaccines by the proportion of the population with naturally acquired immunity. Possible reasons are that the most vulnerable population that may have benefitted from synergistic protection may have already died before these waves or that they were least likely to get infected but most likely to get vaccinated as time went on, boosters included.

In the Omicron wave, the proportion of the population aged 65 years and over that received a booster did not affect COVID-19 death counts, after accounting for seroprevalence in this age group. This suggests that booster doses did not improve upon the effectiveness of the primary vaccine series in preventing severe outcomes, a phenomenon potentially explained by robust cellular immunity elicited by the primary vaccine series. A measurable impact of boosters may have been more likely were variants more closely related to vaccine prototypes circulating during this wave.

Previously, we developed the model to estimate the proportion of the population ever infected based on serology data in New York City and Connecticut [8]. Here, we expand expanded on our previous study and estimated the duration of seropositivity, IFR, and cumulative incidence of SARS-CoV-2 infection as of December 2020 in all 50 states. The average duration of seropositivity could not be estimated in the majority of the states due to either 1) the peak in the seroprevalence in 2020 likely happened before the national serosurveillance started in August 2020, and thus, the whole picture of the timeline of seroreversion was not cauptured, or 2) decline in seroprevalence was never observed. The model we developed works best when there was a single peak in case counts, followed by little infections [8]. New York City and Connecticut were the perfect examples where a single large peak was observed in the spring of 2020 and almost no cases in the summer and early fall of 2020. There were other states that had the similar trend in case counts in 2020, but as the national serosurveillance started in August 2020, the peak in the seroprevalence in many states was missed. Another reason was that the reported seroprevalence never declined in many states, in which case the model estimated that people never returned to seronegative status.

The estimated duration of seropositivity in 2020 in New York and Pennsylvania was consistent with other reports [21]. The estimated duration of seropositivity in this study was longer than the original estimate [8], which is because we had longer duration of seroprevalence data and mortality data, allowing us to observe a longer trajectory of the timeline of antibody waning. We should also note that the average duration of seropositivity was estimated for 2020 based on the seroprevalence data and mortality data in 2020. During this period, reinfection was less common and vaccines were not yet available for the general population. The adjustment of seroprevalence for the timeline of seroreversion at the population level was more important for this reason, compared to the later period of the pandemic where the antibody level was consistently boosted by multiple exposures. The model did not fit the reported seroprevalence data in five states, likely because the assumptions of the model (constant IFR over time) did not hold for these states.

Our study had limitations. The samples collected for the CDC seroprevalence data may not be representative of the general population [14]. We did not have access to individual-level data on infection and vaccine administration, so we could not distinguish groups of people who were infected, vaccinated, and both. All of our data were at the state level. As vaccine coverage varied across geographic area, data at a finer geographic scale might be helpful to identify associations between naturally acquired immunity and vaccine-induced immunity at the population level.

In conclusion, vaccination with the primary series was strongly associated with reduction in COVID-19 mortality at state level, which was sustained through the Alpha wave, Delta wave, and Omicron wave. This effect did not vary by the state-level seroprevalence or estimated proportion of the population ever infected. The understanding of the relationship between vaccine-induced immunity and naturally acquired immunity is critical for post-licensure vaccine evaluation. Ongoing evaluations to monitor mortality in vaccinated populations can guide future policies on boosters and strain changes in the vaccine.

## Data Availability

All data used in this study are publicly available online at the CDC websites cited in the manuscript.

## ACKNOWLEDGEMENT

We would like to thank Dr. Manish Patel at the Centers for Disease Control and Prevention for invaluable feedback and guidance on this study.

## SUPPLEMENTARY TABLES

**Supplementary Table 1.** Results of the mixed-effect linear regression model evaluating the relationship of weekly COVID-19 death counts per 1 million population with coverage of ≥1 dose of COVID-19 vaccine and the seroprevalence in the Alpha wave.

|  | Estimated coefficient (95% confidence interval) |  |
| --- | --- | --- |
|  | With interaction | Without interaction |
| Seroprevalence among people $\geq 65$ years of age (in %) | 0.77 (-0.52, 2.06) | 0.09 (-0.15, 0.33) |
| Coverage of $\geq 1$ dose of COVID-19 vaccines among people $\geq 65$ years of age (in %)* | -0.19 (-0.39, -0.00) | -0.29 (-0.37, -0.22) |
| Interaction of the above two terms | -0.01 (-0.02, 0.01) | NA |

\*Note that the main text reports an estimated change per a 10% increase in coverage.

**Supplementary Table 2.**
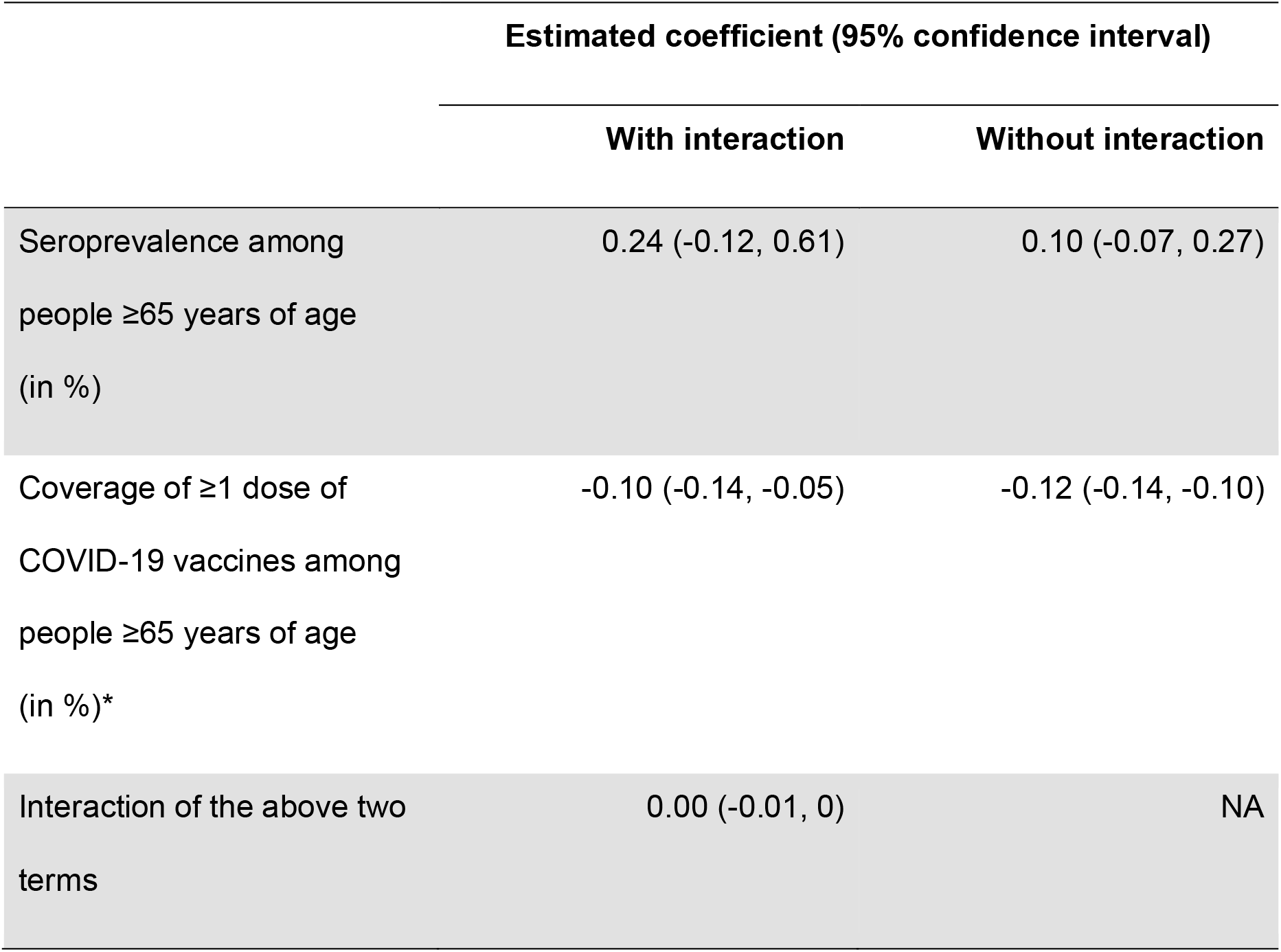
Results of the mixed-effect linear regression model evaluating the relationship of weekly COVID-19 death counts per 1 million population with coverage of ≥1 dose of COVID-19 vaccine and the seroprevalence in the Alpha wave, after incorporating the lags between infection and seroconversion or death.

|  | Estimated coefficient (95% confidence interval) |  |
| --- | --- | --- |
|  | With interaction | Without interaction |
| Seroprevalence among people $\geq 65$ years of age (in %) | 0.24 (-0.12, 0.61) | 0.10 (-0.07, 0.27) |
| Coverage of $\geq 1$ dose of COVID-19 vaccines among people $\geq 65$ years of age (in %)* | -0.10 (-0.14, -0.05) | -0.12 (-0.14, -0.10) |
| Interaction of the above two terms | 0.00 (-0.01, 0) | NA |

**Supplementary Table 3.** Results of the linear regression models evaluating the relationship of total COVID-19 death counts per 1 million population with coverage of ≥1 dose of COVID-19 vaccine and the seroprevalence in the Delta wave.

|  | Estimated coefficient (95% confidence interval) |  |
| --- | --- | --- |
|  | With interaction | Without interaction |
| Seroprevalence among people $\geq 65$ years of age (in %) | -129.25 (-336.53, 78.03) | -3.42 (-16.23, 9.39) |
| Coverage of $\geq 1$ dose of COVID-19 vaccines among people $\geq 65$ years of age (in %)* | -56.62 (-84.78, -28.47) | -42.00 (-56.73, -27.28) |
| Interaction of the above two terms | 1.37 (-0.88, 3.62) | NA |

\*Note that the main text reports an estimated change per a 10% increase in coverage.

**Supplementary Table 4.** Results of the linear regression models evaluating the relationship of total COVID-19 death counts per 1 million population with coverage of ≥1 dose of COVID-19 vaccine and the seroprevalence in the Omicron wave.

|  | Estimated coefficient (95% confidence interval) |  |
| --- | --- | --- |
|  | With interaction | Without interaction |
| Seroprevalence among people $\geq 65$ years of age (in %) | -431.68 (-1937.36, 1073.99) | 8.53 (-0.92, 17.97) |
| Coverage of $\geq 1$ dose of COVID-19 vaccines among people $\geq 65$ years of age (in %)* | -149.06 (-509.93, 211.81) | -44.34 (-88.2, -0.49) |
| Interaction of the above two terms | 4.64 (-11.23, 20.51) | NA |

\*Note that the main text reports an estimated change per a 10% increase in coverage.

**Supplementary Table 5.** Results of the mixed-effect linear regression model evaluating the relationship of weekly COVID-19 death counts per 1 million population with coverage of ≥1 dose of COVID-19 vaccine and the estimated proportion of the population ever infected, using Shioda, *et al*. model [1] in the Alpha wave.

|  | Estimated coefficient (95% confidence interval) |  |
| --- | --- | --- |
|  | With interaction | Without interaction |
| Estimated population ever infected as of December 2020 (in %) | 0.2 (-0.68, 1.09) | 0.07 (-0.09, 0.23) |
| Coverage of $\geq 1$ dose of COVID-19 vaccines among people $\geq 65$ years of age (in %) | -0.24 (-0.44, -0.04) | -0.27 (-0.35, -0.19) |
| Interaction of the above two terms | 0 (-0.01, 0.01) | NA |

[1] Shioda, *et al. Epidemiology* 2021

**Supplementary Table 6.** Results of the mixed-effect linear regression model evaluating the relationship of weekly COVID-19 death counts per 1 million population with vaccine coverage and other estimates of the proportion of the population ever infected [2] in the Alpha wave.

|  | Estimated coefficient (95% confidence interval) |  |
| --- | --- | --- |
|  | With interaction | Without interaction |
| Estimated population ever infected as of December 2020 (in %) | 0.55 (-0.11, 1.21) | 0.09 (-0.03, 0.21) |
| Coverage of $\geq 1$ dose of COVID-19 vaccines among people $\geq 65$ years of age (in %) | -0.14 (-0.35, 0.06) | -0.28 (-0.35, -0.21) |
| Interaction of the above two terms | -0.01 (-0.01, 0.00) | NA |

[2] Chitwood, *et al. medRxiv* 2021

**Supplementary Table 7.** Results of the linear regression models evaluating the relationship of total COVID-19 death counts per 1 million population with coverage of ≥1 dose of COVID-19 vaccine and the estimated population ever infected as of November 2021 [3] in the Omicron wave.

|  | Estimated coefficient (95% confidence interval) |  |
| --- | --- | --- |
|  | With interaction | Without interaction |
| Estimated population ever infected as of November 2021 (in %) | -673.86<br>(-1527.49, 179.77) | 3.85 (-2.18, 9.88) |
| Coverage of $\geq 1$ dose of COVID-19 vaccines among people $\geq 65$ years of age (in %) | -308.52 (-641.4, 24.36) | -46.3 (-88.55, -4.06) |
| Interaction of the above two terms | 7.15 (-1.85, 16.15) | NA |

[3] Barber, *et al. The Lancet* 2022

